# Omicron Impact in India: Analysis of the Ongoing COVID-19 Third Wave Based on Global Data

**DOI:** 10.1101/2022.01.09.22268969

**Authors:** Rajesh Ranjan

**Author notes:** Contributing authors.

## Abstract

The Omicron variant of coronavirus has caused major disruptions world-wide with countries struggling to manage the overwhelming number of infections. Omicron is found to be significantly more transmissible compared to its predecessors and therefore almost every impacted country is exhibiting new infection peaks than seen earlier. In this work, we analyze the global statistics of Omicron-impacted countries including South Africa, the United Kingdom, the United States, France, and Italy to quantitatively estimate the intensity and severity of recent waves. Next, these statistics are used to estimate the impact of Omicron in India, which is experiencing an intense third wave of COVID-19 since 28 Dec., 2021. The rapid surge in the daily number of infections, comparable to the global trends, strongly suggests the dominance of the Omicron variant in infections in India. The logarithmic regression suggests the early growth rate of infections in this wave is nearly four times that in the second wave. Another notable difference in this wave is the relatively concurrent arrival of outbreaks all across the country; the effective reproduction number (***R***_***t***_) although has significant variations among different regions. The test positivity rate (TPR) also displays a rapid growth in the last **10** days in several states. Preliminary estimates with the Susceptible-Infected-Removed (SIR) model suggest that the peak in India to occur in late January 2022 with a caseload exceeding that in the second wave. Although global Omicron trends, as analyzed in this work, suggest a decline in case fatality rate and hospitalizations compared to Delta, a sudden accumulation of active infections can potentially choke the already stressed healthcare infrastructure for the next few weeks.

## 1 Introduction

The world experienced devastating COVID-19 waves in 2021 which were primarily driven by the Delta or B.1.617 variant of the deadly SARS-CoV-2 virus. This variant was not only twice more infectious than its earlier predecessors (Alpha, Beta) but also caused serious illness and causalities [1], especially to those who were not vaccinated. Towards the end of 2021, when the delta variant was continuing to create havoc in several regions of the world, a new variant B.1.1.529 or ‘Omicron’ was detected in South Africa (SA). Soon it was declared as a variant of concern (VOC) by the World Health Organization (WHO) on Nov. 26, 2021, due to several mutations including 15 mutations in the receptor-binding domain (RBD) of spike protein [2] as well as increased transmissibility [3–5]. Omicron quickly replaced the Delta strain in SA to give a new daily peak of more than 37000 cases in a record time of just 20 days compared to a maximum of 26000 per day infections in all the previous waves.

By the end of 2021, Omicron spread to more than 100 countries including the United Kingdom (UK), the United States (US), France, Italy, and India causing new waves. The worldwide daily number of infections shot to about 2.8 million on Jan. 7, 2022, compared to the highest peak of about 0.7 million prior to Omicron. The US alone had reported more than 1 million cases on Jan. 4, 2022. Several studies [2, 3, 6] have raised concern about Omicron evading the antibodies thus increasing the chances of reinfection as well as breakthrough infections. Recent research [7] suggests that three doses of mRNA-based vaccines may also not be sufficient to prevent infection and symptomatic disease with the Omicron variant, whereas another study [8] found the contrary. Despite these conflicting reports, which are preliminary in nature, almost all studies converge on the view that the severity of COVID-19 reduces due to vaccination even with Omicron, and therefore clinical manifestations of the disease could be mild to moderate. This is encouraging news as it may help in the reduction of the risk of hospitalizations and deaths despite a large number of infections. Another silver lining is that some early data suggest that Omicron infections themselves may be milder compared to those caused by earlier variants [9].

The present study concerns the analysis of the global Omicron infection data of most-affected countries to draw insights about the intensity and severity of the new waves that the world is experiencing. Further, these insights are put in the context of the ongoing third wave in India to estimate the possible impact of Coronavirus. This third appears to be driven by the Omicron variant, although genomic surveillance may be necessary to ascertain this. Figure 1 shows different waves of COVID-19 infection that India has observed since the beginning of the pandemic in early 2020. The peak cases in the first and second waves were about 0.1 and 0.4 million respectively. Also, the growth curve in the second wave was much steeper than the first wave. The third wave began on Dec. 28, 2021, nearly six months after the onset of the second wave. At the time of writing the current manuscript, it is about 12 days into the third wave, but daily cases increased from 6,340 on Dec. 27, 2021, to 1,60,610 on Jan. 08, 2022 - a 25 fold increase in 13 days. The curve for the third wave is therefore steeper than the second wave- a characteristic of the outbreaks driven by Omicron as seen in several other countries (e.g. SA, UK, France, Italy, and US).

**Fig. 1:**
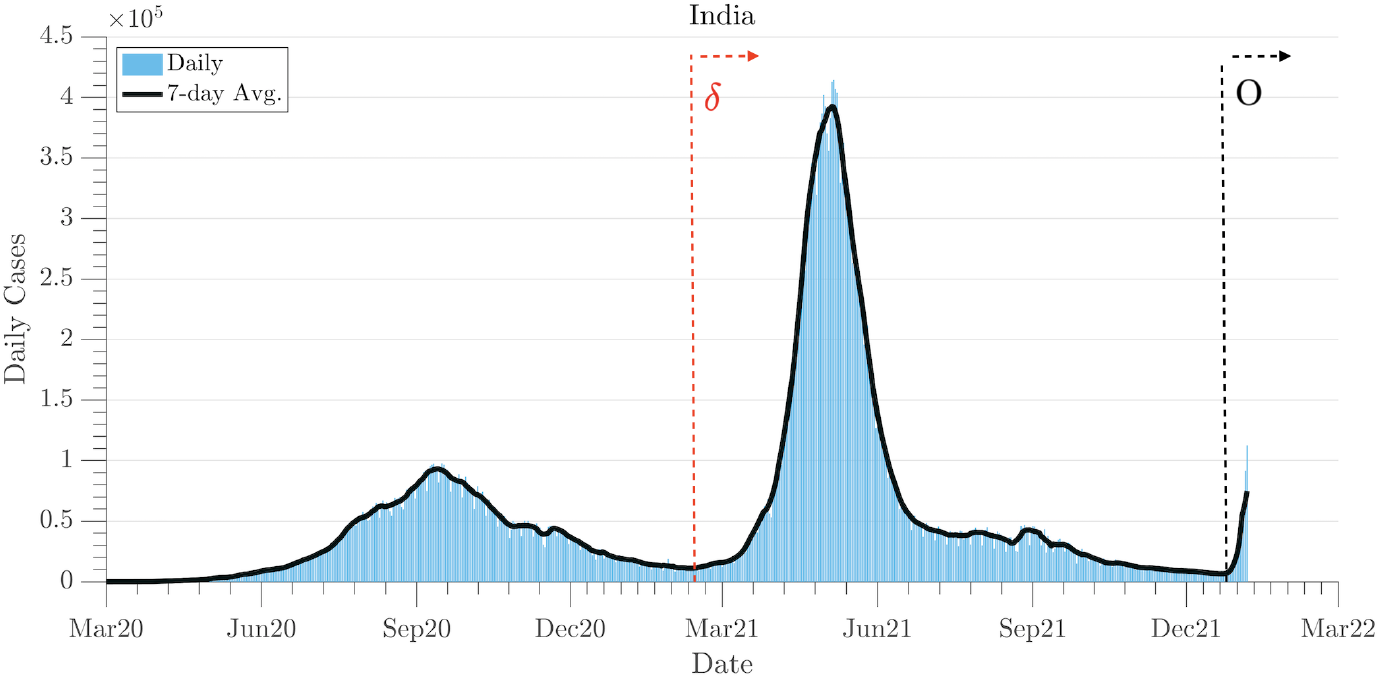
COVID-19 waves in India. The first and second waves as well as the onset of the third wave are shown. Arrows with *δ* and *O* indicate pandemic to be primarily driven by Delta and Omicron variants respectively. The dominance of Omicron in the third wave will be established later.

India has a nearly 1.3 billion population and is 22^*nd*^ most visited nation in the world. Therefore, there is significant interest globally in the possible course and consequences of the current wave. India has recently seen a devastating second wave due to the Delta variant which claimed about 0.33 lives besides tremendous damage to the socio-economic structure. Detailed information about Omicron characteristics such as its pathogenicity, which may be valuable in estimation of possible aftereffects of Omicron infections, is not yet available, and therefore this adds to uncertainty and unpredictability. The present study, therefore, focuses on the characterization of the ongoing third wave: intensity, severity, regional variations, the possible progression of the pandemic, etc., purely from the perspective of global data. The growth curve in the current wave is compared against those of the Omicron-affected countries which are ahead of India in terms of onset. This helps establish whether the current wave is primarily due to the spread of Omicron. Further, the available data is used to estimate key metrics of the wave: exponential growth rate, effective reproduction number [10], test positivity rate (TPR). Regional variations in the wave are also investigated. Finally, an epidemiological model is used to make projections about peak and eventual decay. Throughout the study, the parameters from the third wave are juxtaposed against global data and those from the second wave in India for a critical comparison.

## 2 Dataset

The present study is based on COVID-19 time-series data from the beginning of the pandemic to Jan. 8, 2022. These data are taken from several sources: 1) *ourworldindata* (OWID; [11]) for all the countries, 2) *covid19india*.*org* (COVID19INDIA; [12] and *covid19tracker*.*in* (COVID19TRACKER; [13] for India and her states. OWID curates data from the European Centre for Disease Prevention and Control (ECDC), while COVID19TRACKER/COVID19INDIA compiles data from the official union and local government sources. Unless mentioned otherwise, a 7-day rolling average of daily data was taken for computation of all relevant parameters as well as for the modeling.

## 3 Results

### 3.1 Intensity of wave

We first discuss the intensity of the ongoing third wave in India and compare it to that in Omicron-affected countries. Figure 2 shows the increase in cases in the first 11 days of the Omicron in six countries including India. Since the beginning of the wave is characterized by exponential growth, we plot the number of cumulative infections since the beginning of the Omicron outbreak on a logscale. All the countries exhibit nearly similar growth in the initial days. This trend suggests the predominant SARS CoV-2 Omicron variant accounts for a large proportion of the cases in the current wave in India. Further, it is expected that like SA and most other regions, Omicron will soon replace the Delta variant.

**Fig. 2:**
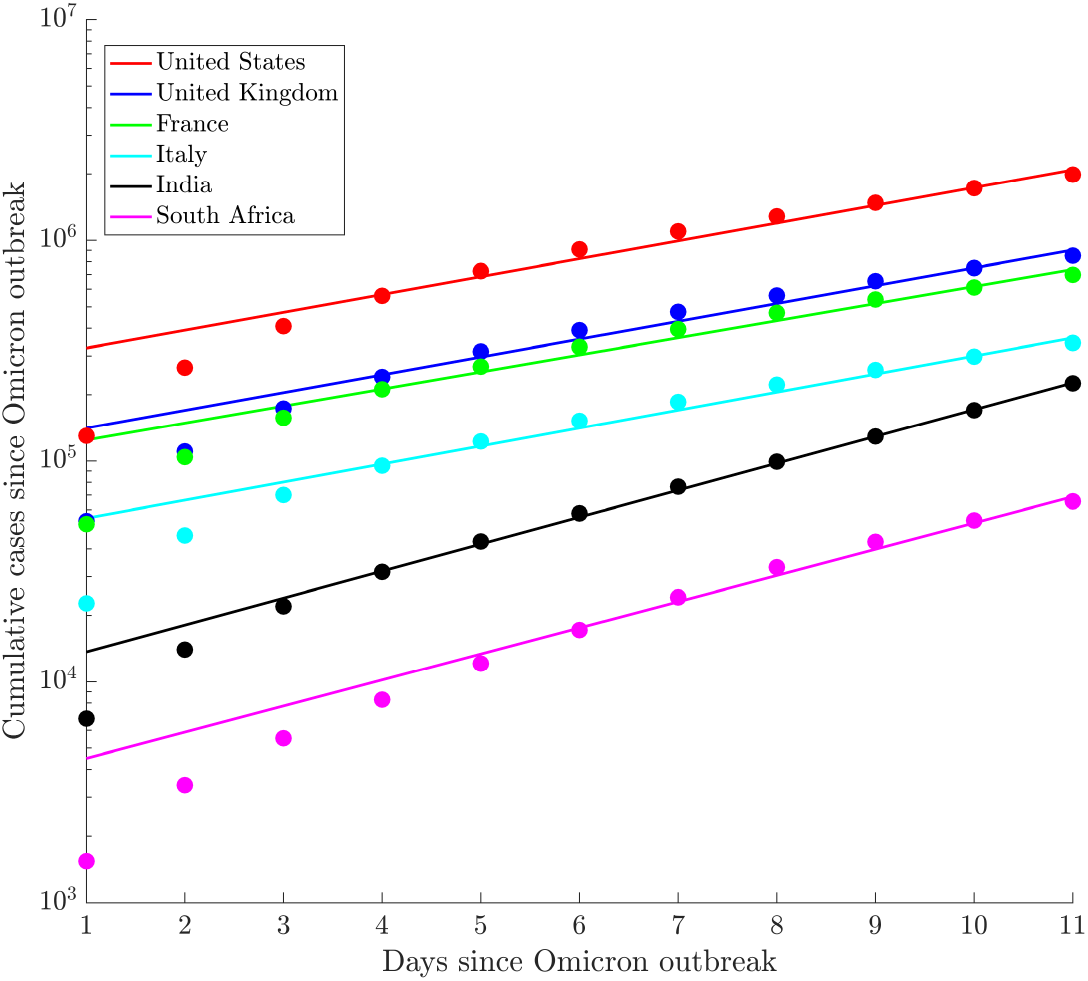
The intensity of the Omicron outbreak in 6 affected countries including India during the first 11 days of the spread. Exponential fits are used to estimate the growth rate. Markers show the actual number of cases, while the lines show the exponential fits. Table 1 presents the exponential factors as obtained from the fit. The onset date of wave for this plot is also included in the table.

To further quantify, we employ exponential fits as shown by lines in Fig. 2. The parameters of these fits are given in Table 1. A key observation is that the growth rate, given by parameter *r* in the fit *I*(*t*) = *I*_0_ exp(*rt*), is nearly similar (about 0.18) in the US, UK, France, and Italy as also seen in Fig. 2. This gives the doubling time as about 3.8 days. India has a slightly higher initial growth rate with *r ≃* 0.28 which is comparable to that in SA. When compared to the second wave in India, as discussed in [14], this factor is nearly four times higher. Based on these rates, doubling time during the exponential growth phase in this wave is about 2.5 days compared to about 10 days in the second wave. Table 1 also lists key goodness of fit parameters. All the fits have a high value of the coefficient of determination (*R*^2^ ≥ 0.97).

**Table 1:**
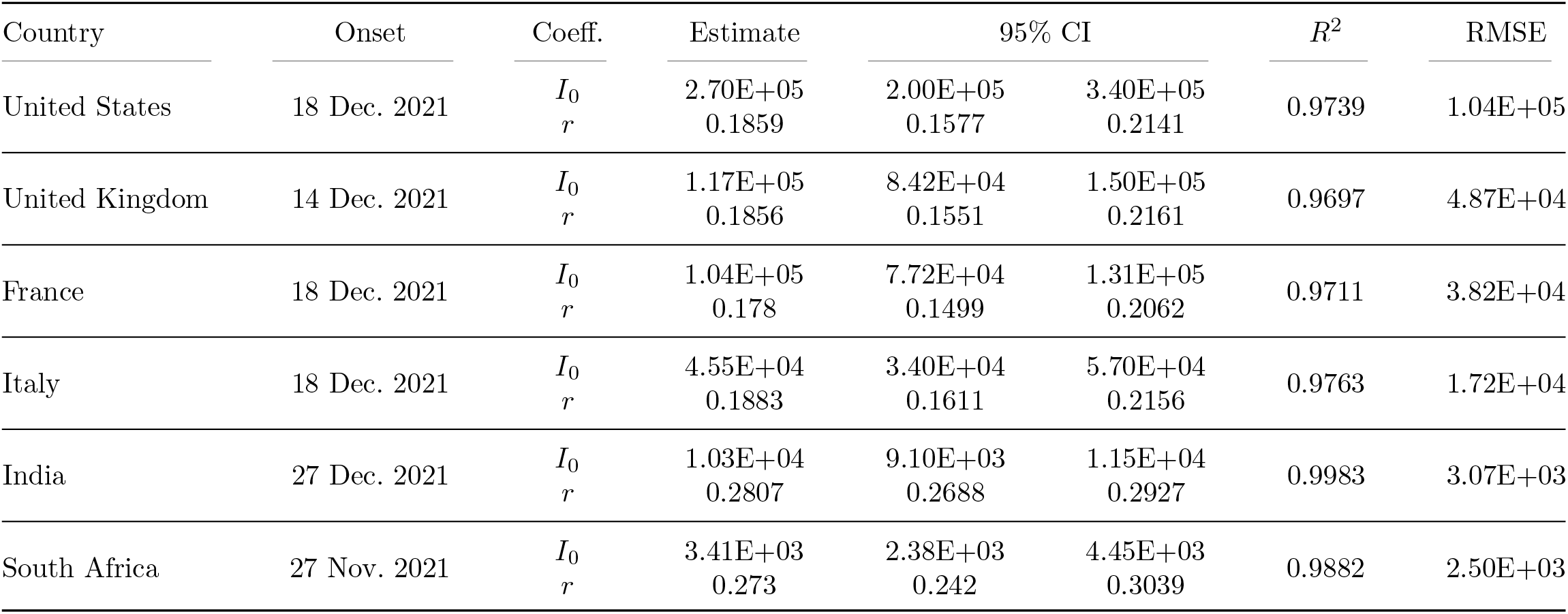
Exponential regression model (*I*(*t*) = *I*_0_ exp(*rt*)) for initial growth in Omicron outbreak in six countries.

### Regional Variations

Next, we discuss the regional variations in the intensity of local outbreaks. India has 28 states, which are administratively governed by locally elected governments of different political parties. Apart from these, there are eight union territories. The list of all the states as well as four large union territories are given in table 2. During the second wave in India (see, Fig. 1), states were separated by a few weeks to months in the onset of the outbreak. For example, Maharashtra showed the signs of a second wave quite early (in mid-February 2021), while in West Bengal and other northeast states the signs were visible only in late March or early April. Figure 3(a) shows the onset of the third wave in India and eight representative states including West Bengal. All the states, except Kerala, show a surge in infections since late December 2021. Kerala is an exception as this state showed a rebound of infections after the second wave (June 2021) when most other states were in the continuous decline phase. Although not shown (discussed later), almost every region in India shows signs of the third wave on January 5, 2021.

**Table 2:** Key metrics of the epidemic spread in Indian states and Union Territories as of Jan. 10, 2022.

| Region | TPR (%) | $R_t$ | Vac. 1 (%)<sup>1</sup> | Vac. 2 (%)<sup>2</sup> |
| --- | --- | --- | --- | --- |
| **India** | 9.3 | 2.33 | 66.0 | 47.1 |
| Andhra Pradesh | 2.8 | 2.13 | 80.8 | 59.6 |
| Arunachal Pradesh | 8.7 | 3.36 | 55.5 | 44.0 |
| Assam | 4.1 | 2.42 | 67.6 | 47.1 |
| Bihar | 2.3 | 3.28 | 50.0 | 35.8 |
| Chandigarh | 15.7 | 3.18 | 89.0 | 67.8 |
| Chhattisgarh | 7.4 | 2.74 | 65.5 | 44.8 |
| New Delhi | 21.2 | 2.63 | 80.0 | 60.0 |
| Goa | 24.2 | 2.26 | 88.3 | 72.7 |
| Gujarat | 7.6 | 2.38 | 73.5 | 63.2 |
| Himachal Pradesh | 9.9 | 2.51 | 84.8 | 75.5 |
| Haryana | 10.5 | 2.79 | 72.4 | 51.7 |
| Jammu and Kashmir | 1.2 | 1.74 | 76.6 | 71.5 |
| Jharkhand | 6.4 | 2.19 | 51.4 | 32.4 |
| Karnataka | 5.8 | 2.77 | 75.8 | 60.6 |
| Kerala | 10.4 | 1.34 | 77.1 | 62.9 |
| Madhya Pradesh | 2.6 | 3.21 | 67.1 | 61.0 |
| Maharashtra | 20.8 | 2.13 | 68.0 | 45.9 |
| Manipur | 3.3 | 1.74 | 43.2 | 32.6 |
| Meghalaya | 2.1 | 2.15 | 38.8 | 27.6 |
| Mizoram | 14.2 | 1.69 | 63.9 | 49.6 |
| Nagaland | 2.6 | 1.86 | 35.3 | 27.0 |
| Odisha | 7.1 | 2.99 | 68.2 | 50.0 |
| Puducherry | 13.7 | 3.4 | 58.7 | 38.7 |
| Punjab | 14.6 | 2.84 | 56.7 | 32.9 |
| Rajasthan | 10.4 | 3.21 | 63.6 | 46.8 |
| Sikkim | 45.3 | 3.16 | 81.8 | 72.7 |
| Tamil Nadu | 8.4 | 2.57 | 68.4 | 46.1 |
| Telangana | 3.3 | 2.15 | 78.4 | 54.1 |
| Tripura | 5.6 | 3.37 | 67.2 | 53.6 |
| Uttar Pradesh | 4.1 | 3.57 | 59.6 | 34.7 |
| Uttarakhand | 7.6 | 2.86 | 74.5 | 59.6 |
| West Bengal | 31.5 | 2.26 | 68.0 | 44.3 |

Note: Vaccination percentage is calculated by considering the entire population in the region. However, currently only 15+ are eligible for vaccination.
<sup>1</sup>Partially vaccinated.
<sup>2</sup>Fully vaccinated.

**Fig. 3:**
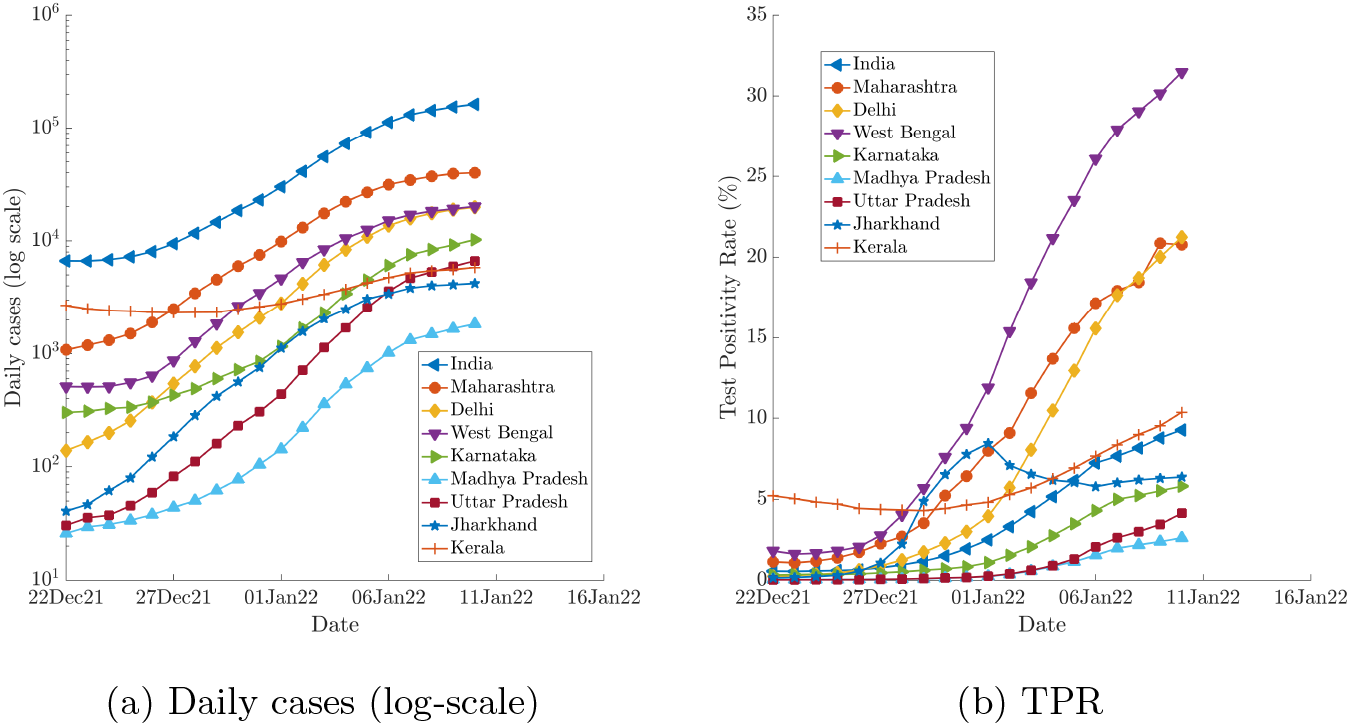
Comparison of COVID-19 spread in India and most-impacted 8 states of India as of January 10, 2022. Figure (a) shows the variations in daily infections in the third wave. Figure (b) exhibits the rapid surge in test positivity rate in this wave.

Another corresponding parameter that characterizes the spread of the outbreak is the test positivity rate (TPR). This is defined as the ratio of number of positive cases per number of tests. Higher TPR indicates a higher level of community transmission. Figure 3(b) shows the TPR for India and eight states discussed earlier. There is a growing trend of TPR in all the states. The TPR in India dramatically increased tenfold in 12 days, from 0.75% on Dec. 27, 2021, to 9.3% on Jan. 10, 2022. As per the latest data, the TPRs for Sikkim and West Bengal are alarmingly high (*>* 30%), while Delhi, Goa, and Maharashtra also have TPR greater than 20%. Table 2 lists the TPR of all the states in India. Punjab(15%), Chandigarh(16%), Mizoram (14%), Rajasthan (10%), Haryana (10%), Puducherry(14%) also have TPRs much higher than the WHO recommended level of 5%. A sudden surge in cases due to the high transmissibility of Omicron has caused a dramatic increase in TPR in every region across the globe it has impacted. For example, the US currently has a TPR of about 25%.

We next discuss another key metric, the effective reproduction number or *R*_*t*_, that represents the average number of infections caused by a single infected individual. Unlike the basic reproduction number or *R*_0_ which is typically fixed for a given outbreak, *R*_*t*_ provides time-accurate variations in the virus transmissibility and hence accounts for temporary interventions, spread, etc. *R*_*t*_ *>* 1 indicates growth in infection. Time-varying *R*_*t*_ is calculated using timeseries data of the infections and generation time distribution [15, 16]. We use the approach developed by Thompson *et al*. [17] and details of the underlying input parameters are given in Ranjan *et al*. [14].

Figure 4 provides the variations of daily cases (incidence) and *R*_*t*_ computed from this data for 8 states; the latest *R*_*t*_ for all the states and populated union territories are listed in Table 2. The effective reproduction number trend typically follows the infection rate curve but reveals transmissibility that may provide insights into the potential future course of the pandemic. A key observation from the figure is the sharp rise in the *R*_*t*_ value in all the states since the onset of the third wave, although the error margin in the computation of *R*_*t*_ at the beginning of a wave may be higher [18]. Kerala, which was an exception after the second wave in terms of continuing growth, also displays the signs of another wave. Most states have also exceeded the peak *R*_*t*_ value in the second wave. These trends suggest that daily cases are going to increase rapidly in the next few days before the arrival of the peak which will be estimated in the next section. Another notable observation, Table 2, is that the third wave has arrived relatively simultaneously in all the states with *R*_*t*_ value greater than the threshold value of 1 for all the regions. This is in contrast to the first and second waves when there was a significant gap of a few weeks to a couple of months in the onset of wave in different states (see figure 4).

**Fig. 4:**
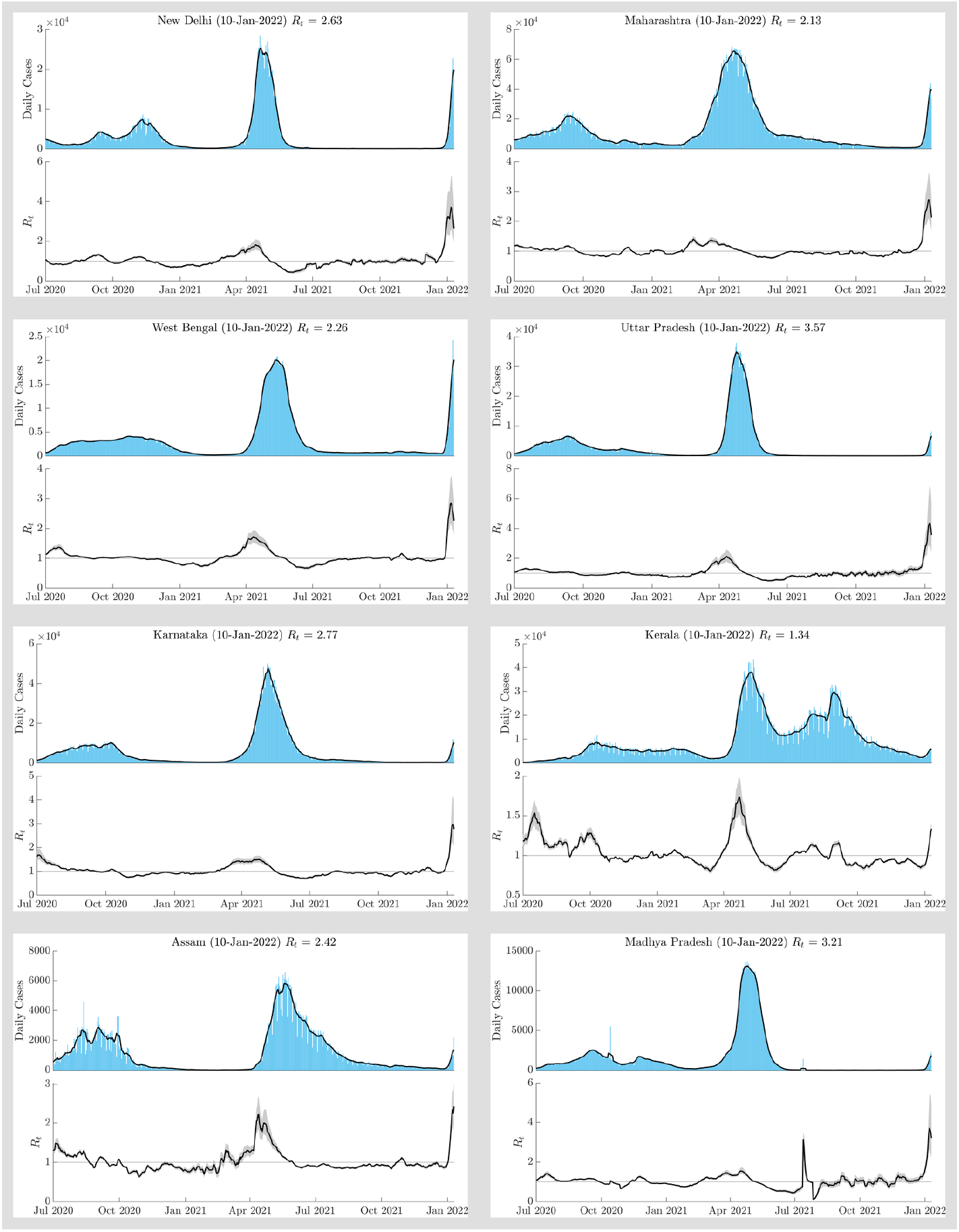
Temporal variations of the effective reproduction number (*R*_*t*_) along with incidence (daily infections) data for 8 key states. The onset of the third wave corresponds with the most recent date when *R*_*t*_ crossed the threshold of 1.0.

### 3.2 Severity of the wave

It has been established by the growth curves that there is a high level of community transmissions in the current wave compared to earlier ones. Now we discuss the potential severity of infections in this wave. Early studies suggest that Omicron infections might be less severe than other variants [9], however more virological data is needed to ascertain this. We rely on time-series data-based approach to gather some insights on the possible impact of Omicron on severe infections and fatality.

Since it has been only two weeks into the third wave, we will employ insights from other Omicron-affected countries to make informed estimates. In order to extrapolate the data from these countries to India, we need to compare the vaccination levels. As alluded earlier, it has been found [7] that Omicron can evade neutralizing antibodies, obtained either from the vaccination or natural infection, and thus there is a greater probability of breakthrough infections. Despite this, vaccinations provide protection from severe infections and hence vaccination levels can have an impact on the severity of Omicron infections.

Despite a large population of 1.33 billion, India currently has about 47% fully vaccinated people. About 0.9 billion in the population (66%) is inoculated with at least one dose. In comparison, the US(74%), the UK(76%), France(79%), Italy(81%) have relatively high vaccination percentages of at least one dose, whereas South Africa (32%) has a low vaccination percentage compared to India. Relatively sharper growth in India and SA in the recent outbreak, as seen in Fig. 2, could be related to relatively lower vaccination levels. The statewise distribution of the partially (Vac. 1) and fully (Vac. 2) vaccinated population as of Jan. 8, 2022, is given in table 2. There is a disparity in vaccination levels among states, with smaller states (Delhi, Goa, Arunchal Pradesh, Sikkim, Haryana, Telangana, Uttarakhand, Jammu and Kashmir, Kerala, Karnataka) generally having higher percentages of vaccinated population, comparable to the US and the UK. Therefore, insights from all these countries can be used to estimate the severity of Omicron impact in India. States with lower vaccination levels, however, may be more vulnerable to relatively severe infections.

Some recent studies [2] suggest a booster dose for further prevention of severe Omicron infection. The US and UK have started booster doses for the eligible population since the third quarter of 2021. Following the onset of the third wave, India started Booster doses for healthcare and frontline workers as well as 60+ populations with co-morbidities. Further, results from South Africa suggest higher hospitalization rates in Omicron among children compared to earlier waves, probably due to lower levels of pre-existing immunity [19]. Until Jan 3, India had only 18+ adults eligible for the vaccination. However, recently India recently started vaccinations for 15+ eligible populations.

Now, we discuss parameters related to the severity of Omicron infections as found in different regions. Figure 5 shows the daily cases, daily case fatality rate (CFR), and weekly case hospitalization rate (CHR) for five Omicron-impacted countries. The case fatality rate is defined as the ratio between confirmed deaths and confirmed cases 10 days earlier as the fatality lags the cases roughly by 10 days [11]. The case hospitalization rate is introduced as a new metric, which is defined as the ratio between weekly new hospitalizations and weekly confirmed cases. Although there is a difference of a few days before Omicron spread into these regions, for simplicity we take Dec. 1, 2021, as the date when trends became visible. The CFRs of all the countries exhibit a decreasing trend in the Omicron wave. SA saw a drastic decay of CFR from about 10% (not visible in fig.) on Nov. 26, 2021, to less than 1% on Jan. 28, 2021. Also, in the US, the UK, France, and Italy, the CFR is less than 1% based on the latest data. Although it remains to be seen if the trend persists. In comparison, CFR was very high in the US, Italy, and France in the period July to August 2021, a few weeks from the onset of a new wave due to the Delta variant.

**Fig. 5:**
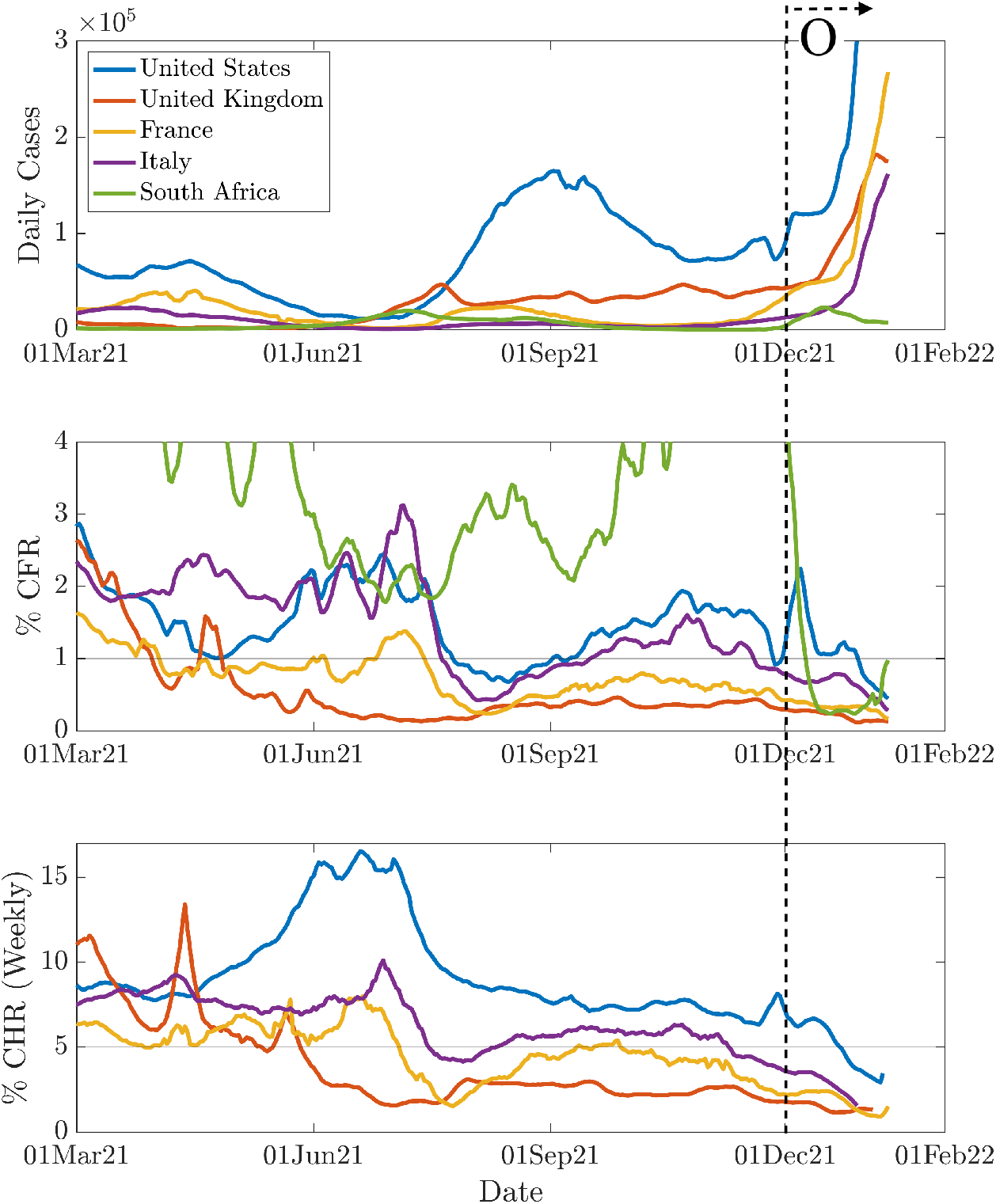
Temporal variations of daily cases, Case fatality rate (CFR), and Case Hospitalization rate (wherever available) in five Omicron-impacted countries. The most recent wave with Omicron is marked with a right arrow on the top right. Note that, y-axis in top panels are truncated to focus on the regions of interest.

Next, we discuss the hospitalization rate from the countries where the weekly hospital admission data due to COVID-19 is available. The CHR curve is generally at a peak during the onset of the Delta wave (June-July 2021). During the most recent wave due to Omicron, the CHR curve shows a declining trend. The UK, France, and Italy show less than 3 hospital admissions per 100 infections in the most recent week. The US has slightly higher than 5% CHR; during the Delta wave, the CHR peaked at more than 15%. In the last week, both France and the US have shown a mild increase in CHR. This relatively low CHR in Omicron is also expected to be characteristics of the third wave in India. The will also consequently improve the case recovery rate (CRR), defined as the ratio between number of recoveries and number of infections, thereby reducing the active caseload. These factors overall suggest that the severity of the third wave will be lesser than the second wave despite high community transmission.

### 3.3 Modeling and Future Estimates

After characterization, we employ an epidemiological model on the third wave data to make useful estimates on the possible progression of the pandemic. Specifically, we employ the dynamical Susceptible-Infected-Remove (SIR) model [20], which takes the time-series infection data to estimate the peak as well as the eventual decline of the pandemic. This model along with its variations (SEIR, SIRD) has been used in many regions for the modeling of COVID-19 behavior [21]. The author has successfully used the SIR model to make projections of the course of pandemics in the second COVID-19 wave [14]. Both the peak caseload as well as date were reasonably well predicted by this model in advance of about a month from the peak. Obtaining reliable estimates for the third wave is more challenging because of the very sharp rise of the wave and expected early arrival of peak (based on observations from SA and UK) and thus leaving a very small window size for the input data. However, it is hoped that the estimates will be useful in policy-making decisions if not quantitatively precise.

We employ the MATLAB implementation of the SIR model as developed by [22]. This model is tweaked for the Indian data as delineated in [14]. Figure 6 shows the predictions for India using the SIR model along with other key characterizing metrics discussed earlier. The input data used for the projection is taken between 27 Dec. 2021 and 5 Jan. 2022. Statewise and districtwise estimates for most affected regions are available on our website www.forecast-covid19.org.

**Fig. 6:**
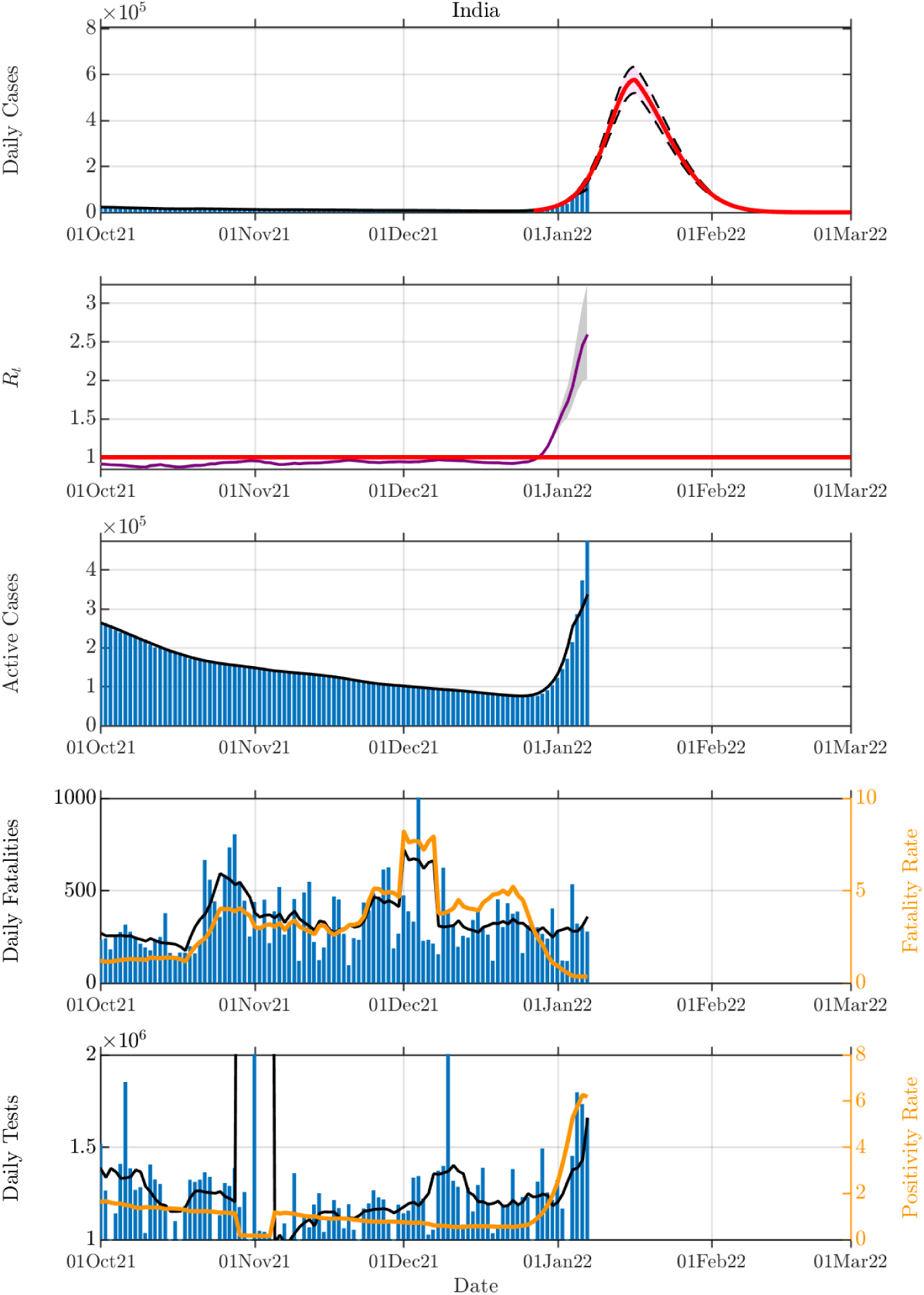
Predictions of the COVID-19 third wave in India using classical Susceptible-Infected-Recovered (SIR) model.

According to the model (top panel in Fig. 6) based on about 10 days of data, the third wave is expected to peak around the third week of January. Individual states are also expected to peak around this time with a lag or lead of one week. Therefore, the period of the third wave could be much shorter than the earlier waves. The second wave took about three months (mid-Feb to mid-May) to the peak. Despite a short period, the peak caseload in the current wave (*>* 0.4 million) could be higher than the second wave. This rapid surge in numbers could stress the healthcare systems in the coming weeks, despite the expected low CHR.

The model also suggests a rapid decline in the number of cases following the peak. As observed earlier, the case recovery rate (CRR) in the current wave is expected to be higher; this will ease the burden of active caseload compared to the second wave despite the higher number of daily infections. The peak in active cases is expected to come about two weeks following the peak in daily infections. The peak in daily fatalities should follow the peak in active cases. The rapid increase in the number of cases needs to match with a proportional increase in the daily testing capacity in order to keep the test positivity rate (TPR) to the recommended level of below 5%. Further, since the current wave has arrived relatively simultaneously in all the states, this may restrict the mobilization of patients and resources across the states to some level.

Despite indications from the model about rapid growth and rapid decline of cases along with fewer hospitalizations and deaths compared to the second wave, a major concern is the emergence of a new variant due to high community transmission. This may alter the projected course of the pandemic. Further, any change in the testing protocol can also significantly change the predicted caseload. The projections also need to be updated with the availability of the new data.

## 4 Summary

In this work, the global Omicron infection data in the six most impacted countries - the US, the UK, India, Italy, France, and Italy - are analyzed to draw key insights. The daily infection curves indicate rapid growth in all these countries with a doubling time of 2.5-4 days, significantly lesser than Delta waves. Despite high transmissibility, the data indicates a decline in case hospitalization and case fatality rates during Omicron in all the countries analyzed. For example, in the US, the case hospitalization rate reduced from a maximum of 15% during the Delta wave to less than 4% during the ongoing Omicron outbreak. This could be a combined effect of relatively milder strain as well as protective effects of vaccinations. With specific to India, currently experiencing an intense third wave, the study has the following observations:

- The intensity of infection and transmission in the third wave is significantly higher than the second wave.
- The early trend of daily cases in India compares well with the early trend data of other Omicron-affected countries (SA, US, UK, Italy, France), thus indicating that the third wave is predominantly driven by Omicron.
- Signs of the third wave are visible across every state and union territory in India. This contrasts with the second wave when the onsets were offset by a few weeks to months.
- Based on data from other Omicron-affected countries, despite high transmission, hospitalizations and fatalities per infection is expected to be lower.
- Preliminary analysis with the SIR model suggests a peak within a month of the onset of the third wave. The decline is also expected to be faster than the second wave.

## Data Availability

All data produced are available online at https://ourworldindata.org/ and https://covid19tracker.in/

https://ourworldindata.org/

https://covid19tracker.in/

https://covid19-forecast.org/

## 5 Acknowledgment

The author would like to thank Prof. Mahendra K. Verma for the insightful discussions. Acknowledgments are also due to Dr. Sudheendra Rao N R, Dr. Deepti Chugh for discussions at various points. Mr. Aryan Sharma and Mr. Rahul Garg provided help with plots and other material on our COVID-19 website.

## Statements & Declarations

## Data availability

All the data that support the findings of this study are publicly available.

